# Initial evaluation of a mobile SARS-CoV-2 RT-LAMP testing strategy

**DOI:** 10.1101/2020.07.28.20164038

**Authors:** Christina M. Newman, Mitchell D. Ramuta, Matthew T. McLaughlin, Roger W. Wiseman, Julie A. Karl, Dawn M. Dudley, Miranda R. Stauss, Robert J. Maddox, Andrea M. Weiler, Mason I. Bliss, Katrina N. Fauser, Luis A. Haddock, Cecilia G. Shortreed, Amelia K. Haj, Molly A. Accola, Anna S. Heffron, Hailey E. Bussan, Matthew R. Reynolds, Olivia E. Harwood, Ryan V. Moriarty, Laurel M. Stewart, Chelsea M. Crooks, Trent M. Prall, Emma K. Neumann, Elizabeth D. Somsen, Corrie B. Burmeister, Kristi L. Hall, William M. Rehrauer, Thomas C. Friedrich, Shelby L. O’Connor, David H. O’Connor

## Abstract

Severe acute respiratory syndrome coronavirus 2 (SARS-CoV-2) control in the United States remains hampered, in part, by testing limitations. We evaluated a simple, outdoor, mobile, colorimetric reverse transcription loop-mediated isothermal amplification (RT-LAMP) assay workflow where self-collected saliva is tested for SARS-CoV-2 RNA. From July 16 to November 19, 2020, 4,704 surveillance samples were collected from volunteers and tested for SARS-CoV-2 at 5 sites. A total of 21 samples tested positive for SARS-CoV-2 by RT-LAMP; 12 were confirmed positive by subsequent quantitative reverse-transcription polymerase chain reaction (qRT-PCR) testing, while 8 were negative for SARS-CoV-2 RNA, and 1 could not be confirmed because the donor did not consent to further molecular testing. We estimated the RT-LAMP assay’s false-negative rate from July 16 to September 17, 2020 by pooling residual heat-inactivated saliva that was unambiguously negative by RT-LAMP into groups of 6 or less and testing for SARS-CoV-2 RNA by qRT-PCR. We observed a 98.8% concordance between the RT-LAMP and qRT-PCR assays, with only 5 of 421 RT-LAMP negative pools (2,493 samples) testing positive in the more sensitive qRT-PCR assay. Overall, we demonstrate a rapid testing method that can be implemented outside the traditional laboratory setting by individuals with basic molecular biology skills and can effectively identify asymptomatic individuals who would not typically meet the criteria for symptom-based testing modalities.

## Introduction

More than 340,000,000 severe acute respiratory syndrome coronavirus 2 (SARS-CoV-2) diagnostic tests have been performed in the United States as of February 22, 2021, yet it is estimated that 80-95% of infected individuals are not tested ^1,2^. The availability of diagnostic testing for population surveillance around the United States has been limited because of testing supply shortages and guidelines set by public health officials ^3,4^. Multiple studies have shown that asymptomatic and presymptomatic individuals infected with SARS-CoV-2 can be as infectious as symptomatic individuals ^5–9^, with recent estimates of up to 59% of transmission coming from asymptomatic or presymptomatic individuals ^10^. Virological assessments of SARS-CoV-2-positive individuals and coronavirus disease 2019 (COVID-19) patients further support the reports of asymptomatic transmission, identifying no significant differences in viral loads found in the upper respiratory tracts of asymptomatic and symptomatic individuals ^5,7,11–13^. Furthermore, Arons et al. (2020) demonstrated that positive viral cultures can be isolated from presymptomatic patients up to 6 days before the onset of symptoms ^5^.

Delays in reporting test results can prevent timely isolation of infected individuals. Most current testing programs fail to identify and efficiently notify infected individuals. Since transmission can occur before symptoms manifest, reporting delays create a major barrier to safely returning to workplaces and schools ^14^. Therefore, there remains an urgent need for rapid tests that identify presymptomatic and asymptomatic individuals while conserving diagnostic testing reagents. Non-diagnostic point-of-care (POC) testing, used in conjunction with the current clinical diagnostic testing regimen, may improve our ability to identify infectious individuals and limit their exposure to others while they are most contagious and conserve clinical diagnostic tests for those who require confirmatory testing. Incorporating active surveillance using POC tests as part of mitigation strategies for reopening K-12 schools could play an integral role in reducing SARS-CoV-2 transmission among students, teachers and staff members, families, and the surrounding community ^15,16^.

Loop-mediated isothermal amplification (LAMP) is a low-cost method for rapid target-specific detection of nucleic acids ^17^. LAMP has long been used as an alternative to gold-standard quantitative reverse transcription polymerase chain reaction (qRT-PCR) to surveil populations for a variety of pathogens, especially in resource-limited settings ^18–22^. Reverse transcription LAMP (RT-LAMP) assays have recently been developed for rapid SARS-CoV-2 testing ^23–29^. RT-LAMP is an appealing candidate for POC SARS-CoV-2 testing because it is inexpensive, circumvents supply shortages by relying on different reagents than current diagnostic tests, requires minimal sample processing, and can be deployed outside of traditional laboratory settings. Recently, a number of studies have shown the correlation between the presence of virus in saliva and nasopharyngeal swabs, demonstrating that saliva specimens are a valid and reliable alternative to nasopharyngeal swab specimens for SARS-CoV-2 testing ^30–35^. Saliva specimen self-collection is noninvasive, can be done at home, does not require swabs or personal protective equipment, and limits direct contact between test operators and testing populations. Here we describe our experience implementing a simple, rapid-turnaround, mobile, non-diagnostic SARS-CoV-2 testing workflow combining self-collected saliva and RT-LAMP in volunteers without symptoms of SARS-CoV-2 infection. Individuals were strongly encouraged to isolate and obtain follow-up diagnostic testing after receiving a positive result by RT-LAMP. This addresses a key knowledge gap of how on-site RT-LAMP testing performs in real-world conditions, since virtually all previous studies have only evaluated SARS-CoV-2 RT-LAMP in well-equipped molecular biology laboratories.

## Materials and Methods

### POC testing sites

To begin operating voluntary POC testing, we developed a system of color-coded storage bins for equipment and supplies, as well as assembled folding tables, chairs, extension cords, and coolers that could be easily decontaminated and packed to fit in a Dodge Caravan (FCA US LLC., Auburn Hills, MI) or other, similarly sized minivan for transportation between testing sites and our base laboratory facility. On July 16, 2020, we launched our first mobile POC testing sites which ultimately expanded over 18 weeks to include two workplaces, two K-12 schools, and an athletics program (Suppl. Table 1). With the exception of the athletics program, sites were initially outdoors, sometimes under an overhang, but otherwise open to the environment. The athletics site was a climate-controlled, indoor practice field. At all sites, equipment and reagents were transported by minivan and surfaces were disinfected during assembly, breakdown, and frequently throughout testing. Participant consenting and volunteer sample collection were performed on-site but separated from the sample preparation and assay areas (most commonly on the other side of the building). In an effort to limit contamination, each assay area was set up with three separate folding tables: (1) sample heat-inactivation and preparation, (2) preparation of RT-LAMP reagents and assay set-up, and (3) RT-LAMP incubation and imaging. Individuals responsible for sample inactivation and performing assays wore appropriate personal protective equipment (PPE) including N95 face masks, face shields or safety glasses, disposable lab coats, and double gloves. In anticipation of wet and cold fall weather, by September 2020, assay workspaces were transitioned to biosafety hoods in a vacant indoor laboratory space for several POC testing locations. In October 2020, we received IRB approval for obtaining consent for repeat SARS-CoV-2 testing. This allowed us to transition away from consenting participants at each testing time point and instead allowed each enrolled participant to consent once regardless of the number of times they supplied a sample. Following reports that SARS-CoV-2 RNA is stable in saliva at room temperature for prolonged periods ^36^, we also transitioned away from in-person sample collection at some of the testing sites and instead distributed self-collection take-home kits for drop off at designated locations for same day processing.

**Table 1.**
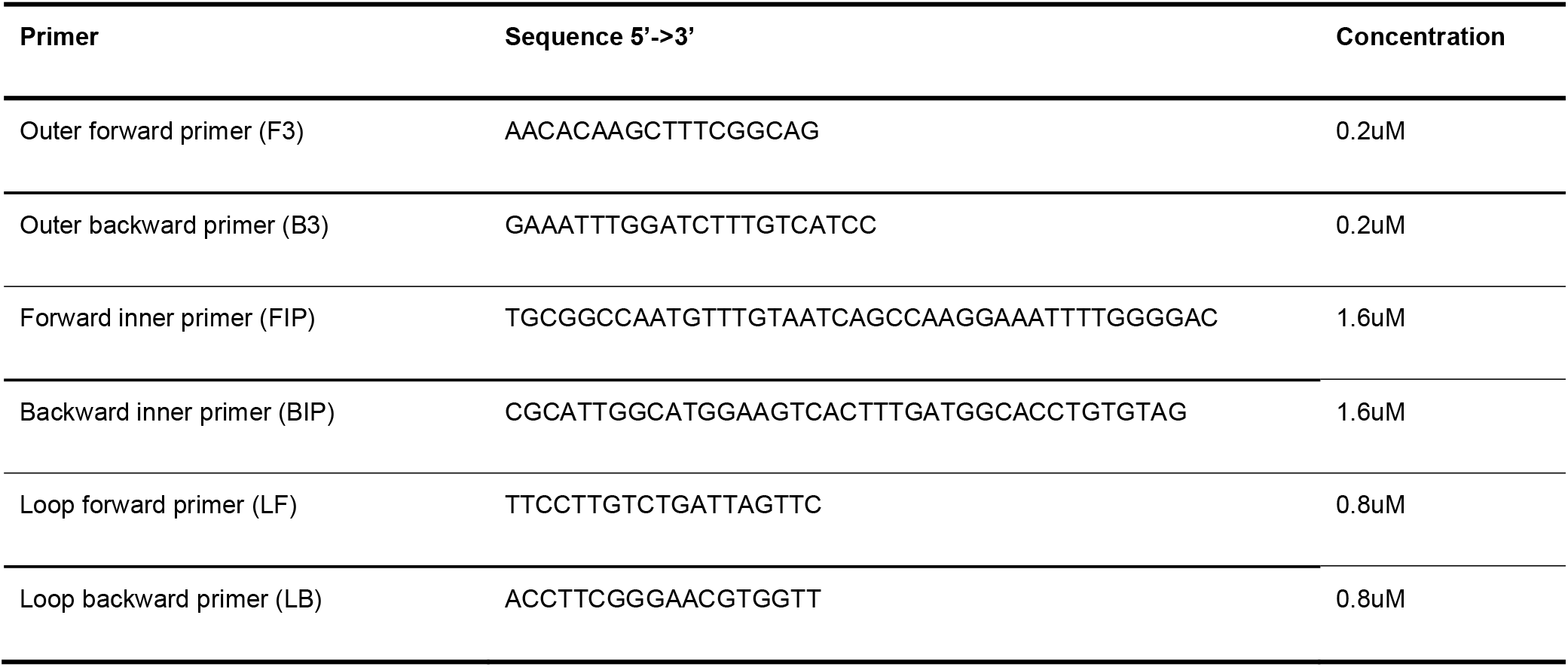
RT-LAMP N-gene primers

### Sample collection and preparation

We obtained approval from the University of Wisconsin-Madison Institutional Review Board (#2020-0855 and #2020-1142). Participants were advised to avoid eating, or drinking anything except for water, for 30 minutes prior to providing a sample. After providing informed consent, volunteers self-collected at least 50 µl of saliva in a 1.5 ml “safe-lock” microcentrifuge tube using a 1000 µl unfiltered pipette tip to funnel the specimen into the tube. Each volunteer disinfected the outside of the tube with a pre-moistened disinfectant wipe. Samples collected in-person were typically processed within 3 hours of collection through our RT-LAMP mobile testing workflow, while samples collected using take-home kits were typically processed within 30 hours (Figure 1). Samples were first incubated in a heat block at 65°C for 30 minutes to inactivate SARS-CoV-2 ^37^ and then incubated in another preset heat block at 98°C for 3 minutes to improve nucleic acid detection and inactivate salivary enzymes ^38^. The inactivated saliva was then centrifuged for 2 minutes in a benchtop microcentrifuge. Fifty microliters of the saliva supernatant were then added to 50 µl of 1x phosphate buffered saline, pH 7.4 (1x PBS).

**Figure 1:**
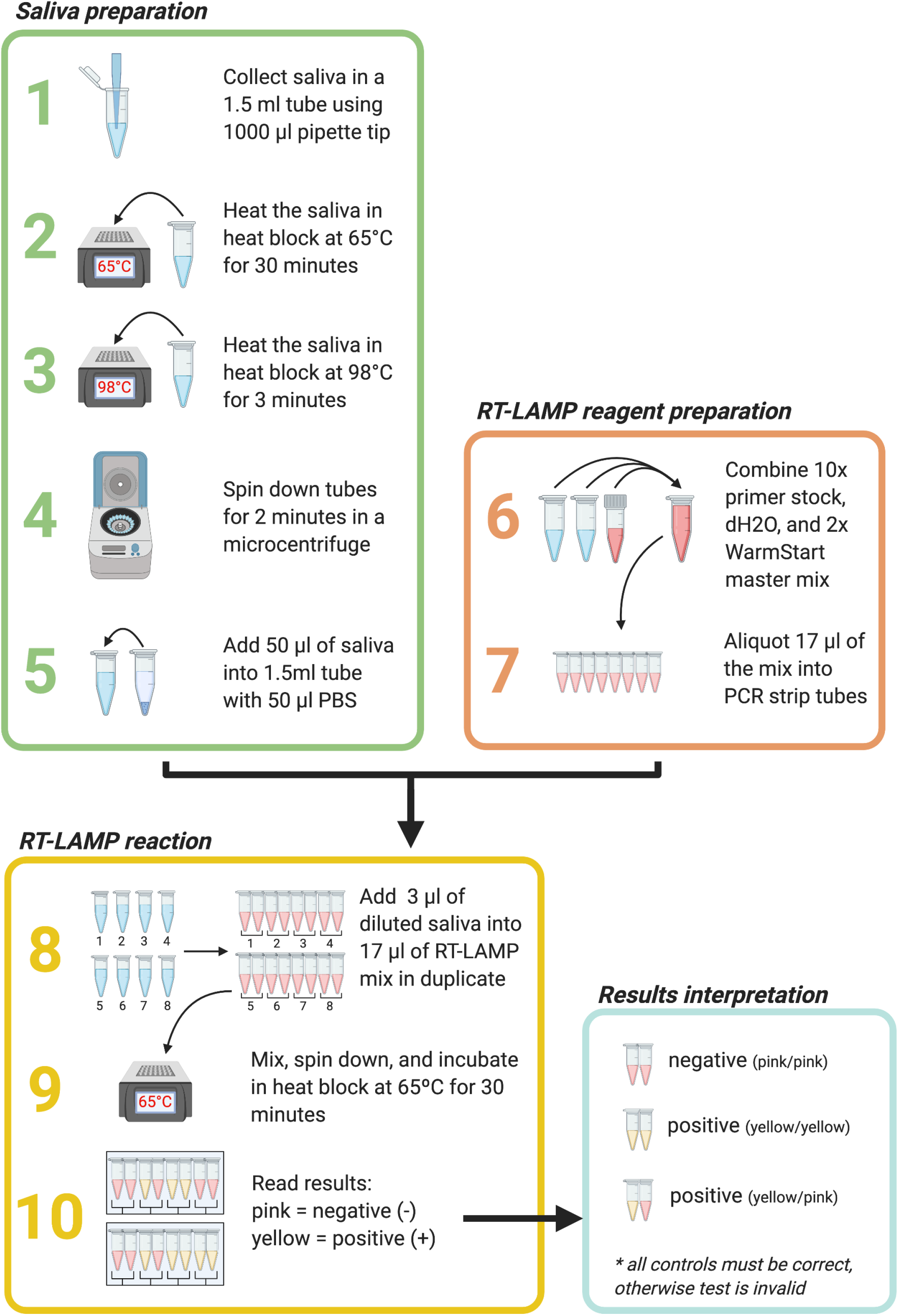
Point-of-care RT-LAMP SARS-CoV-2 testing workflow. **Steps 1-5.** Saliva sample preparation. **Steps 6-7**. RT-LAMP reagent preparation. **Steps 8-10**. RT-LAMP reactions and results interpretation. A reaction color change from pink/orange to yellow after 30 minutes in at least 1 of 2 sample replicates was scored as positive. Figure was created using BioRender.com.

### RT-LAMP reactions

Three microliters of the saliva/PBS mixture for each sample was added in duplicate to 17 µl of a colorimetric RT-LAMP reaction mix containing WarmStart colorimetric LAMP mastermix (NEB, catalogue# M1800), water, and a set of six SARS-CoV-2-specific RT-LAMP primers designed against the N gene ^38^. The SARS-CoV-2 RT-LAMP primer set was previously designed by Broughton et al. and is currently used in an FDA emergency use authorized (EUA) COVID-19 test by Color Genomics (Table 1) ^39,40^. Reactions were incubated for 30 minutes at 65°C. A smartphone or tablet was used to record images of each reaction before (time = 0) and after the incubation period (time = 30). A color change from pink/orange to yellow in at least 1 of 2 replicates was scored relative to gamma-irradiated SARS-CoV-2 (irSARS-CoV-2, BEI Resources, Manassas, VA) that was directly added to RT-LAMP reactions as a positive control in each batch of reactions at concentrations ranging from 220-3,333 copies/µl (2.2×10^5^ - 3.33×10^6^ copies/ml). irSARS-CoV-2 was diluted and aliquoted as ready-to-run positive control standards and stored at -80°C. On the day of testing, the positive controls were removed from the freezer and stored on ice at POC sites. Individuals whose samples were recorded as potentially positive for SARS-CoV-2 by RT-LAMP were contacted by an infectious disease clinician in accordance with the IRB protocol and urged to obtain a clinical diagnostic test to confirm findings and self-isolate in accordance with public health recommendations.

### Limit of detection (LOD) estimation using contrived saliva samples

To estimate the limit of detection of the RT-LAMP assay, contrived positive saliva samples were prepared by adding irSARS-CoV-2 diluted from 1×10^4^-10 copies/µl (1×10^7^-1×10^4^copies/ml) or from 5×10^4^-50 copies/µl (5×10^7^-5×10^4^ copies/ml) directly into unaltered saliva collected from a total of 25 SARS-CoV-2-negative individuals. Dilutions were based on two independent, in-house qRT-PCR experiments showing that the ir-SARS-CoV-2 stock concentration ranged from 7.89×10^6^ - 8.23×10^6^ copies/µl (7.89×10^9^ - 8.23×10^9^ copies/ml). In two RT-LAMP experiments, four serial dilutions of irSARS-CoV-2 were prepared for each saliva sample in duplicate. RT-LAMP reactions were set up as described previously. Negative controls consisting of 1x PBS and positive controls consisting of 1×10^4^ copies/µl (1×10^7^ copies/ml) irSARS-CoV-2 in water were also prepared in duplicate. Reactions were called positive if a color change from pre-amplification to post-amplification occurred in at least 1 of 2 replicates that was consistent with that of the positive controls.

### Limit of detection (LOD) estimation using clinical samples

De-identified discard saliva samples from 38 SARS-CoV-2-positive patients were provided by the University of Wisconsin Hospitals and Clinics (UWHC) for evaluation of RT-LAMP performance with known positive saliva samples. Clinical saliva samples were originally collected and stored at 4°C for up to 4 weeks prior to assessment by RT-LAMP. Additional 10-fold and 100-fold dilutions were prepared for 13 of the samples in saliva collected from a negative volunteer. Clinical samples and dilutions were prepared as described previously except that 20-50 µl of heat-inactivated sample, dependent on total sample volume, was added to an equal volume of 1x PBS in a clean 1.5 ml screw-top tube and pipetted gently to mix. For each sample, 3 µl was then added to duplicate colorimetric RT-LAMP reactions. Negative and positive control reactions (described previously) were also prepared in duplicate except that saliva collected from a negative volunteer was used as the negative control for these reactions. RT-LAMP reactions were prepared and images collected as described previously.

### Quantitative RT-PCR

#### POC samples

We measured vRNA concentration using sensitive qRT-PCR in a subset of the inactivated saliva samples described above after initial evaluation using RT-LAMP. Saliva samples that were negative for SARS-CoV-2 by RT-LAMP were pooled into groups of 6 or fewer for qRT-PCR to balance cost effectiveness with reasonable estimated detection sensitivity. Ten additional, individual RT-LAMP-negative samples were submitted as negative controls alongside samples identified as positive by RT-LAMP. Saliva samples that were identified as positive for SARS-CoV-2 by RT-LAMP were tested by qRT-PCR individually to estimate our POC LOD. RNA was isolated from up to 150 µl saliva and combined with an equivalent volume of nuclease-free water using the Viral Total Nucleic Acid kit for the Maxwell RSC instrument (Promega, Madison, WI) following the manufacturer’s instructions. Viral load quantification was performed using a sensitive qRT-PCR assay developed by the CDC to detect SARS-CoV-2 (specifically the N1 assay) and commercially available from IDT (Coralville, IA). The assay was run on a LightCycler 96 or LC480 instrument (Roche, Indianapolis, IN) using the Taqman Fast Virus 1-step Master Mix enzyme (Thermo Fisher, Waltham, MA). The limit of detection of this assay is estimated to be 0.2 genome equivalents/µl (200 genome equivalents/ml) saliva. To determine the vRNA load, samples were interpolated onto a standard curve consisting of serial 10-fold dilutions of *in vitro* transcribed SARS-CoV-2 N gene RNA kindly provided by Nathan Grubaugh (Yale University) and described by Dudley et al. ^35^.

#### Clinical samples

qRT-PCR was performed using the conditions described above for each of the 38 SARS-CoV-2 positive saliva samples individually; however, sample volume limitations required that for some samples, only 100 µl saliva was combined with 100 µl of nuclease-free water prior to RNA isolation. In addition, sample UWHC3 contained a lower volume than the remaining 37 samples so 50 µl saliva was combined with 50 µl nuclease-free water and used for RNA isolation as described previously. Viral loads in copies per microliter and corresponding cycle threshold numbers (Ct) are reported in Table 2.

**Table 2.** RT-LAMP evaluation of SARS-CoV-2 positive clinical saliva samples.

| Sample | Ct (N1 assay) | Positive by RT-LAMP | vRNA load (copies/μl) | Sample | Ct (N1 assay) | Positive by RT-LAMP | vRNA load (copies/μl) |
| --- | --- | --- | --- | --- | --- | --- | --- |
| UWHC1 | 27.65 | 0/2 | $3.25 \times 10^2$ | UWHC20 | 25.80 | 2/2 | $9.48 \times 10^2$ |
| UWHC2 | 32.7 | 0/2 | 10.9 | UWHC21 | 20.18 | 2/2 | $4.40 \times 10^4$ |
| UWHC3 | 20.98 | 2/2 | $5.17 \times 10^4$ | UWHC22 | 28.92 | 0/2 | $1.13 \times 10^2$ |
| UWHC4 | 24.07 | 2/2 | $3.57 \times 10^3$ | UWHC23 | 21.26 | 2/2 | $2.10 \times 10^4$ |
| UWHC5 | 26.53 | 2/2 | $6.81 \times 10^2$ | UWHC24 | 29.92 | 0/2 | 57.2 |
| UWHC6 | 30.85 | 1/2 | 37.4 | UWHC25 | 36.71 | 0/2 | 0.796* |
| UWHC7 | 36.96 | 0/2 | 0.701 | UWHC26 | 25.96 | 2/2 | $1.31 \times 10^2$ |
| UWHC8 | 26.28 | 1/2 | $8.10 \times 10^2$ | UWHC27 | 29.99 | 0/2 | 54.1 |
| UWHC9 | 37.59 | 0/2 | 0.402 | UWHC28 | 24.34 | 2/2 | $2.58 \times 10^3$ |
| UWHC10 | 24.01 | 2/2 | $3.72 \times 10^3$ | UWHC29 | 20.55 | 2/2 | $4.72 \times 10^4$ |
| UWHC11 | 22.39 | 2/2 | $1.10 \times 10^4$ | UWHC30 | 33.18 | 0/2 | 7.89 |
| UWHC12 | 35.46 | 0/2 | 1.75 | UWHC31 | 22.87 | 2/2 | $9.57 \times 10^3$ |
| UWHC13 | 36.09 | 0/2 | 1.14 | UWHC32 | 23.07 | 2/2 | $8.33 \times 10^3$ |
| UWHC14 | 23.11 | 2/2 | $5.96 \times 10^3$ | UWHC33 | 26.85 | 2/2 | $6.20 \times 10^2$ |
| UWHC15 | 23.38 | 2/2 | $4.95 \times 10^3$ | UWHC34 | 20.33 | 0/2 | $5.49 \times 10^4$ |
| UWHC16 | 33.86 | 0/2 | 3.99 | UWHC35 | 23 | 2/2 | $8.88 \times 10^3$ |
| UWHC17 | n/a | 0/2 | 0 | UWHC36 | 32.26 | 0/2 | 14.9* |
| UWHC18 | 23.02 | 2/2 | $6.34 \times 10^3$ | UWHC37 | 33.94 | 0/2 | 4.33 |
| UWHC19 | 37.31 | 0/2 | 0.612 | UWHC38 | 25.96 | 2/2 | $1.74 \times 10^3$ |

\*Sample only positive in one qRT-PCR replicate.

## Results

### LOD estimation using contrived saliva samples

We assessed the LOD for minimally processed saliva samples collected from 25 volunteers over two RT-LAMP experiments using irSARS-CoV-2 spiked into negative saliva samples (Figure 2A and 2B). In our first experiment (S1-S3), we detected irSARS-CoV-2 in at least 1 of 2 replicates at 1×10^2^ copies/µl (1×10^5^ copies/ml) in all 3 samples (Figure 2A). In our second experiment (S4-S25), we detected irSARS-CoV-2 by RT-LAMP in 2/2 replicates at 5×10^4^ copies/µl (5×10^7^ copies/ml) for 95% of samples, at 5×10^3^ copies/µl (5×10^6^ copies/ml) for 62% of samples, and at 500 copies/µl (5×10^5^ copies/ml) for 10% of samples. When we included samples called positive in at least 1 of 2 replicates (see Methods), the percentage of contrived samples positive by RT-LAMP at each of the aforementioned dilutions were 100%, 90%, and 33.3% respectively (Figure 2B). One sample was omitted from the analysis because it turned yellow before the RT-LAMP reaction incubation began and was therefore uninterpretable. Because in POC testing we defined a positive RT-LAMP result as an observed post-incubation color change to yellow in at least 1 replicate, these results suggested that our 90% LOD is between 1×10^2^ and 5×10^3^ copies/µl (1×10^5^ -5×10^6^ copies/ml).

**Figure 2:**
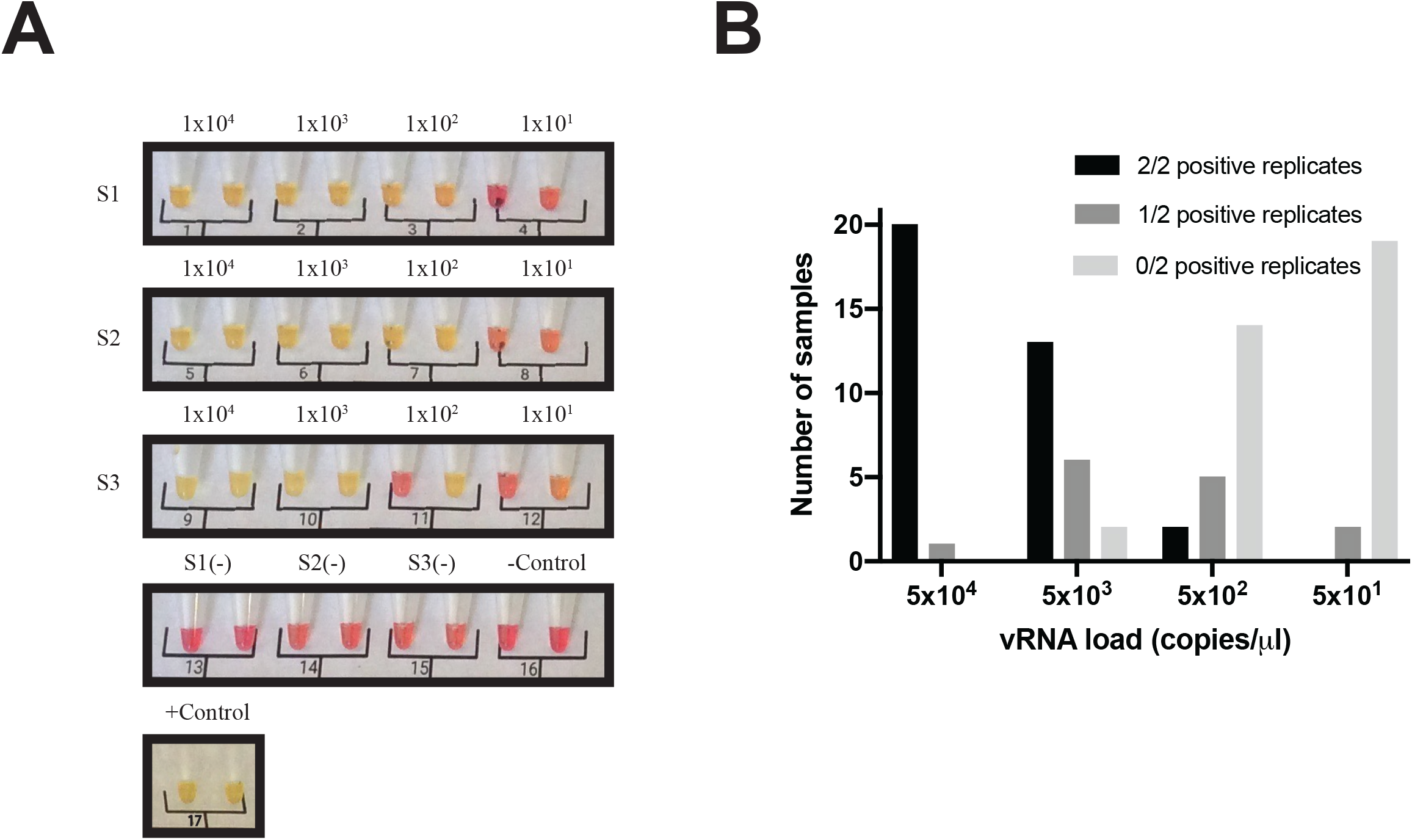
Detection of SARS-CoV-2 in contrived saliva samples by direct RT-LAMP. **A**. Initial limit of detection (LOD) assessment with contrived saliva samples from 3 volunteers (S1, S2, S3). RT-LAMP reactions determined to be negative are pink and those determined to be positive are yellow. Quantitative RT-PCR vRNA loads are presented as copies/µl above the replicates for each sample. **B**. Bar graph showing an expanded assessment of RT-LAMP LOD for 22 additional contrived saliva samples (S4-S25). Gamma-irradiated SARS-CoV-2 (irSARS-CoV-2) vRNA load is shown as copies/µl on the x-axis, number of samples positive in 2 (black), 1 (dark gray), or 0 (light gray) replicates is shown on the y-axis.

### LOD estimation using clinical samples

To assess the performance of SARS-CoV-2 RT-LAMP in known SARS-CoV-2 positive saliva samples as opposed to contrived positive samples, we acquired deidentified, discarded saliva samples collected from 38 patients with laboratory confirmed SARS-CoV-2 from UWHC. Nineteen of 38 undiluted saliva samples were positive for SARS-CoV-2 in 2/2 replicates by RT-LAMP (Figure 3; Table 2). Two additional samples were positive in 1 of 2 replicates. Quantitative RT-PCR data showed that the viral RNA (vRNA) loads of the positive samples ranged from 131 copies/µl to 5.7×10^4^ copies/µl (1.31×10^5^- 5.71×10^7^ copies/ml) which was consistent with our LOD range for contrived samples (Table 3). Furthermore, for the 13 samples diluted 10-fold and 100-fold, detection decreased with increasing dilution factor (Table 4).

**Table 3.** RT-LAMP results for 10- and 100-fold dilutions of 13 SARS-CoV-2-positive samples from UWHC.

| Sample | 1:10 dilution result | 1:100 dilution result | Undiluted vRNA load (copies/μl) |
| --- | --- | --- | --- |
| UWHC1 | 1/2 | 0/2 | $3.25 \times 10^2$ |
| UWHC2 | 0/2 | 0/2 | 10.9 |
| UWHC3 | 2/2 | 2/2 | $5.17 \times 10^4$ |
| UWHC4 | 2/2 | 2/2 | $3.57 \times 10^3$ |
| UWHC5 | 1/2 | 0/2 | $6.81 \times 10^2$ |
| UWHC6 | 0/2 | 0/2 | 37.4 |
| UWHC7 | 0/2 | 0/2 | 0.701 |
| UWHC8 | 1/2 | 0/2 | $8.10 \times 10^2$ |
| UWHC9 | 0/2 | 0/2 | 0.402 |
| UWHC10 | 2/2 | 0/2 | $3.72 \times 10^3$ |
| UWHC11 | 2/2 | 1/2 | $1.10 \times 10^4$ |
| UWHC12 | 0/2 | 0/2 | 1.75 |
| UWHC13 | 0/2 | 0/2 | 1.14 |

**Table 4.**
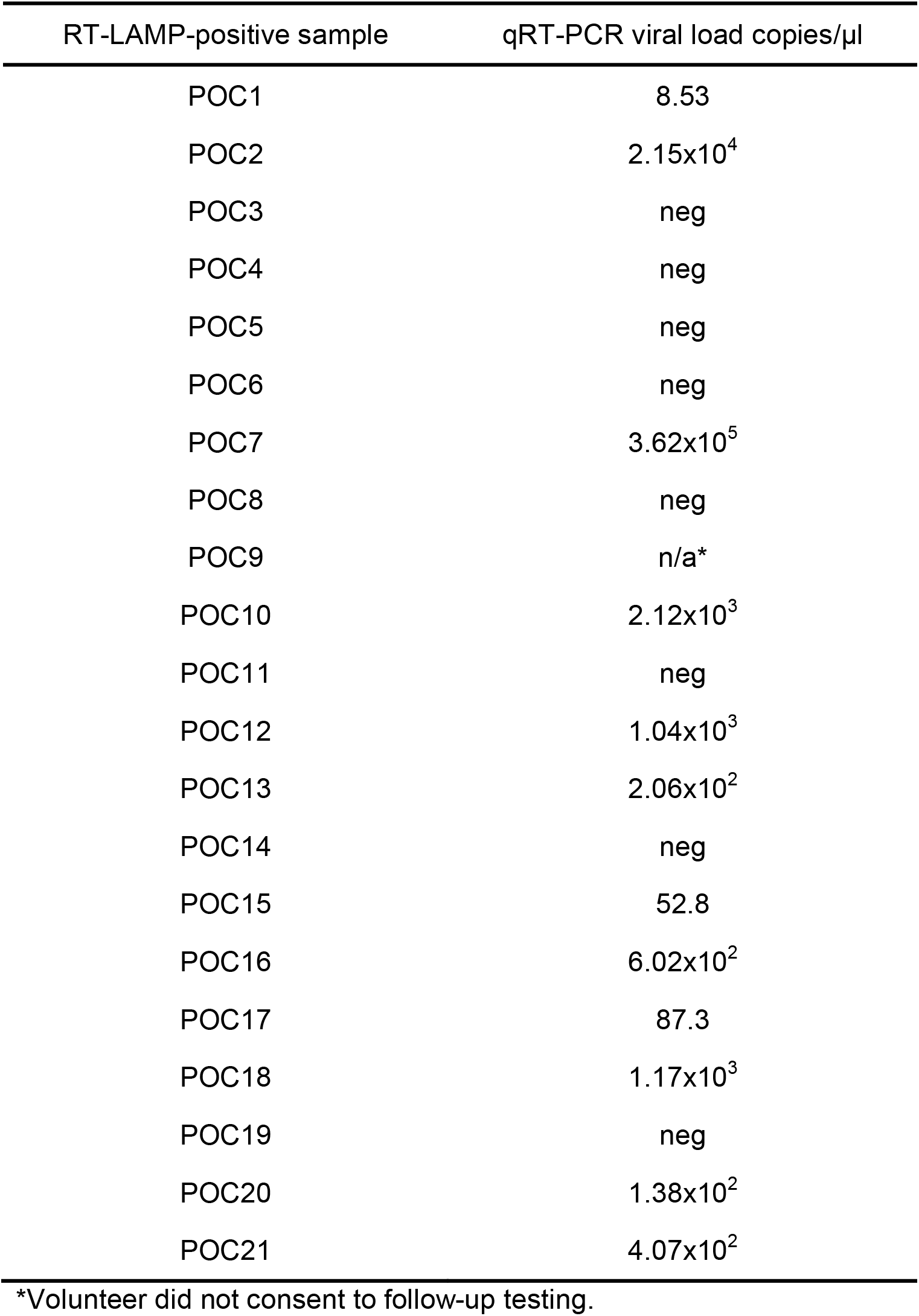
Samples identified as potentially positive for SARS-CoV-2 by RT-LAMP during point-of-need testing.

**Figure 3:**
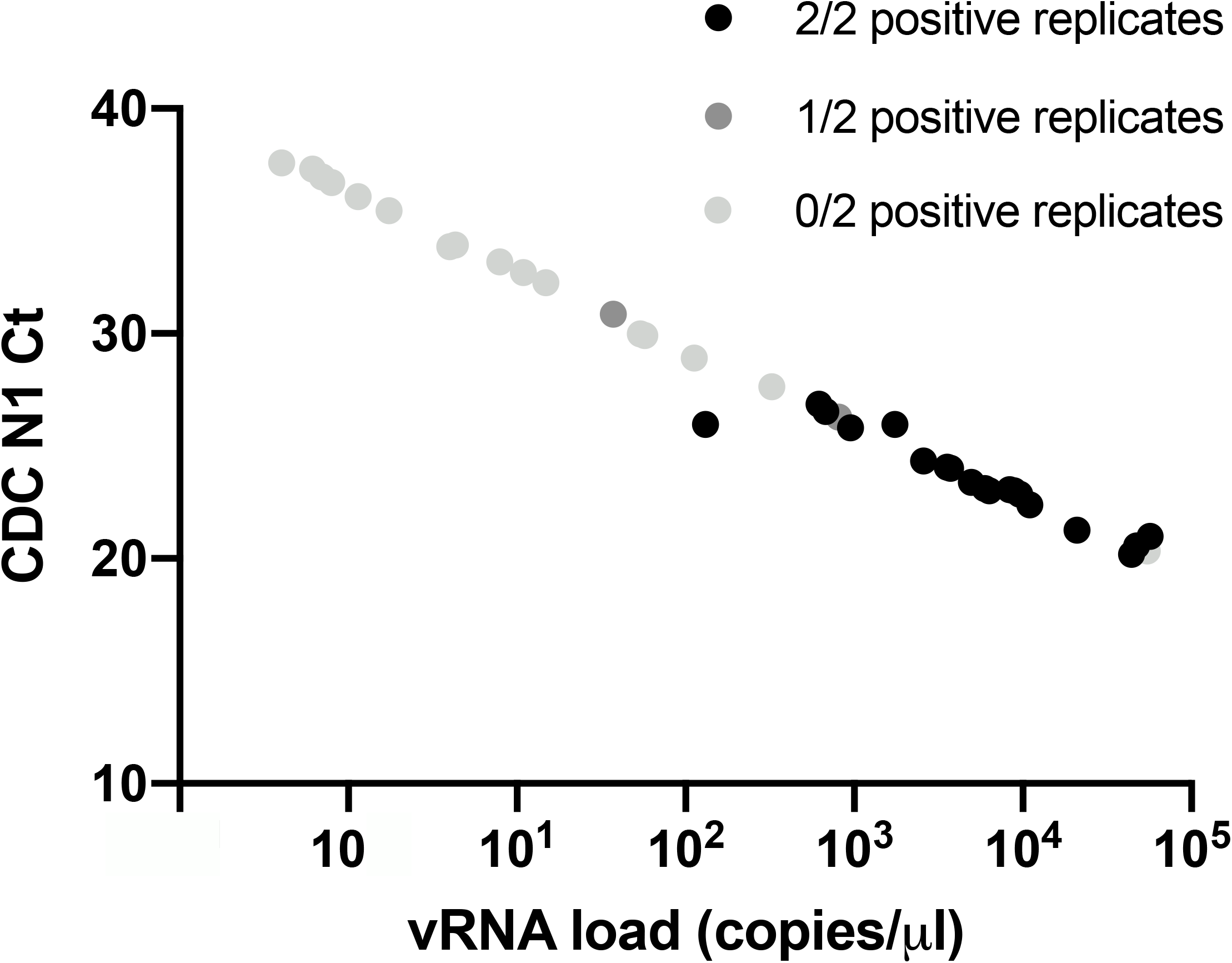
Detection of SARS-CoV-2 in 38 clinical saliva specimens by direct RT-LAMP. The vRNA load of each clinical sample is plotted on the x-axis relative to the in-house CDC N1 qRT-PCR assay cycle threshold (Ct) on the y-axis. Black, dark gray, and light gray indicate 2, 1, and 0 positive replicates respectively.

### POC SARS-CoV-2 RT-LAMP testing

From July 16 to November 19, 2020, SARS-CoV-2 RT-LAMP was used to test a total of 4,704 samples collected from 5 locations. Participants were enrolled into the study regardless of their SARS-CoV-2 symptom status on the day of testing. Seventy-one percent of the samples were obtained from individuals at two research facilities, 11% from two K-12 schools, and 18% from an athletics program (Supplemental Table 1). A total of 21 samples were identified as positive for SARS-CoV-2 by RT-LAMP based on a colorimetric change from pink/orange to yellow in at least 1 of 2 sample replicates. Similar to our experience with our contrived LOD samples, about 0.40% (19/4,704) of samples collected during POC testing exhibited a color change to yellow prior to RT-LAMP assay amplification and were therefore uninterpretable. Follow up qRT-PCR testing was conducted on each sample that appeared positive after the 30 minute amplification reaction throughout the study to determine vRNA load. Twelve of the 21 samples called positive in RT-LAMP had detectable SARS-CoV-2 RNA by qRT-PCR. Viral RNA loads of these samples ranged from 8.58 copies/µl to 3.62×10^5^ copies/µl (8.58×10^3^ copies/ml-3.62×10^8^ copies/ml) with a median of 504.5 copies/µl (5.04×10^5^ copies/ml) (Table 4). Eight of the saliva samples identified as positive by RT-LAMP were negative by qRT-PCR, suggesting that they were false-positive RT-LAMP results. One RT-LAMP-positive sample was not tested by qRT-PCR because the participant did not consent to additional molecular testing. For volunteers who consented to additional research testing from July 16 to September 17, qRT-PCR testing was conducted for pools of 6 or fewer for all residual, heat-inactivated samples that appeared unambiguously negative by RT-LAMP. A total of 421 RT-LAMP-negative pools (2,493 samples) were tested to estimate the number of SARS-CoV-2-positive samples missed by RT-LAMP. Quantitative RT-PCR detected SARS-CoV-2 nucleic acids in 5 pools of RT-LAMP-negative samples. Four out of five of the positive pools contained levels of SARS-CoV-2 that were below the estimated LOD range for RT-LAMP using crude samples with vRNA load estimates of 0.236, 0.444, 0.460, 37.5, and 142 copies/µl (236, 444, 460, 3.75×10^4^, and 1.42×10^5^ copies/ml). Taken together, the low prevalence of SARS-CoV-2 in our volunteer testing population (0.36%, including RT-LAMP-negative, qRT-PCR-positive pools) and the low vRNA load of pools positive by follow-up qRT-PCR, suggest that these 5 pools likely contained only a single positive sample each and suggests a false-negative rate of 0.02% (5/2,493) (Table 4).

## Discussion

Strategic surveillance testing of asymptomatic individuals has been suggested as an important mitigation strategy for places at high risk for close contact, indoor SARS-CoV-2 transmission: schools, workplaces, places of worship, and prisons, among others. Decentralized, mobile RT-LAMP-based POC testing workflows can provide same-day results which can enable people with potential SARS-CoV-2 infections to quickly self-isolate and then obtain confirmatory diagnostic testing. The low per-test cost (approximately $7 per sample tested in duplicate) allows for repeated testing to identify incident infections and reduce the duration of a potentially infected individual’s exposure to others. While RT-LAMP is not as sensitive as diagnostic qRT-PCR tests in laboratory testing, qRT-PCR tests require centralized labs, which in turn leads to lengthy turnaround times. Over a period of 18 weeks, we performed 4,704 SARS-CoV-2 tests across 5 sites using a simple, saliva-based, direct RT-LAMP assay. This work demonstrates the scalability of decentralized, mobile RT-LAMP-based testing and addresses a key knowledge gap of how POC RT-LAMP testing performs outside of well-equipped molecular biology laboratories.

Our initial experiments using direct RT-LAMP with contrived saliva samples from a total of 25 donors demonstrated a LOD that ranged from 1×10^2^ copies/µl (100% in at least 1 replicate for S1-S3) to 5×10^3^ copies/µl (90% in at least one replicate for S4-S25). Taken together, these data suggest that the actual LOD for RT-LAMP without RNA isolation may be dependent on the individual sample due to heterogeneity of saliva pH and composition ^41–43^. Overall, the RT-LAMP results for 38 clinical saliva samples obtained from SARS-CoV-2-positive individuals at the UWHC, were consistent with those for the contrived samples. We recognize that more clinical samples are required for a comprehensive clinical validation, but the LOD observed in clinical samples is further supported by the low vRNA loads obtained from qRT-PCR-confirmed SARS-CoV-2-positive samples identified in our volunteer population (Table 4). The performance of our RT-LAMP POC testing workflow demonstrates that inexpensive, mobile testing can be successfully performed outdoors or in other non-traditional laboratory settings to identify SARS-CoV-2-positive individuals regardless of whether or not symptoms are present. Our observed SARS-CoV-2 RT-LAMP positivity rate was 0.25% (12/4,704) for samples confirmed by follow-up qRT-PCR. Interestingly, the positivity rate of 0.25% in our volunteer population was lower than expected given the disease activity in our region during this period of time was listed as “critically high”, particularly between September 1 and November 19, 2020 when the county had a 5.42% positivity rate (19,031 positive tests out of 350,722) ^44,5^. The low positivity rate in our volunteer population may be partly explained by the fact that 71% of tested saliva specimens came from two research facilities where mask wearing and physical distancing guidelines were implemented early in the pandemic and followed relatively stringently (Supplemental Table 1). Volunteers for nonsymptomatic research testing might also have a different risk profile from the overall population.

Potential drawbacks of colorimetric RT-LAMP-based surveillance for SARS-CoV-2 as described here include the fact that minimally-processed saliva can result in variable reaction color change without the presence of the target RNA. However, modifications of RT-LAMP-based SARS-CoV-2 assays to reduce saliva sample variability, improve result ambiguity, and increase throughput have recently been reported elsewhere and may improve the implementation of RT-LAMP-based assays for POC use ^46–50^. In addition, we relied on a manual RT-LAMP format during POC testing. Reading assays “by eye” inevitably results in a somewhat subjective determination of positives. Reducing false-positive results in our POC volunteer populations required consistent use of duplicate reactions for each individual, which reduced assay throughput and increased the per-sample cost. Furthermore, the testing landscape changed dramatically during the months we performed RT-LAMP testing. The first non-instrumented antigen test, the Abbott BinaxNOW COVID-19 Ag CARD, received FDA EUA approval in the United States on August 26, 2020 ^51^. While the sensitivity of RT-LAMP is broadly comparable to the Abbott BinaxNOW antigen test (reported as 1.6×10^4^ - 4.3×10^4^ vRNA copies; Ct 30.3-28.8), because the former is technically straightforward and can be used as a SARS-CoV-2 diagnostic at testing sites operating under a Clinical Laboratory Improvement Amendments (CLIA) waiver, it is likely a better choice for rapid turnaround, on-site testing in most circumstances ^52^. However, even with the existence of antigen tests, RT-LAMP surveillance programs still have a place as part of a comprehensive SARS-CoV-2 risk mitigation strategy, especially in areas where access to antigen tests is limited.

There are advantages to continuing saliva-based RT-LAMP surveillance testing. Importantly, the supply of diagnostic antigen tests remains tightly constrained, and in the United States, these tests are available only through government contracts. Widespread testing of individuals without symptoms with such a scarce resource may not be a wise use of these limited tests. Furthermore, recent studies have shown that antigen test performance may differ between asymptomatic and symptomatic populations. Compared to qRT-PCR, the sensitivity of FDA-approved antigen tests, BinaxNOW and the Quidel Sofia SARS Antigen Fluorescent Immunoassay, were 35% and 41% in asymptomatic individuals and 64% and 80% in symptomatic individuals, respectively ^53,54^. BinaxNOW is currently only approved for use in symptomatic individuals, within 7 days of symptom onset, and samples are required to be tested within an hour of collection ^55^. In contrast, RT-LAMP reagents do not require a government contract and can be acquired readily from commercial and non-commercial sources, and they can also be used more flexibly for surveillance purposes because RT-LAMP is not limited to use in symptomatic individuals ^56^. Additionally, user acceptance of testing may also favor saliva-based RT-LAMP as it is less invasive than nasal swab-based tests. While an individual BinaxNOW test is rapid, performing several tests during a single day could cumulatively take as long as processing a batch of tests by RT-LAMP. For these reasons, RT-LAMP may still be the preferred testing method to incorporate into a local program. In Madison, WI, two local schools have implemented RT-LAMP surveillance programs modeled on the program described here. Implementation of each program required approximately 50 hours of hands-on training by our group. School staff were trained in adherence to regulations pertaining to non-diagnostic testing and to competently perform RT-LAMP assays. Each school also needed time and resources to acquire the modest lab infrastructure necessary to perform RT-LAMP. In addition, a larger saliva-based RT-LAMP surveillance program has been successfully implemented in school districts in the greater Chicago suburbs ^57,58^.

A looming question for both RT-LAMP and antigen testing programs is whether the real-world effectiveness of frequently testing individuals without symptoms mirrors the theoretical benefits. Several important considerations that we factored into the design of RT-LAMP testing remain true: nonsymptomatic individuals account for up to 59% of all transmission (24% asymptomatic and 35% presymptomatic); low-sensitivity tests are able to effectively identify those with high levels of virus shedding, and individuals with high viral loads are likely to be responsible for a significant fraction of onward community transmission; and the duration of peak infectiousness is short, so lengthy lags in reporting test results could miss a critical window of high transmissibility ^10,59^. Conversely, high-quality, exceptionally well-resourced testing programs such as those at the White House and among intercollegiate athletic programs have failed to stop outbreaks ^60^. The latter deserves special note: outbreaks in these programs occurred in spite of 100% adherence to daily testing. Data from daily sampling of individuals with incident SARS-CoV-2 infection suggests that the mean duration of time from infection to peak viral shedding is approximately three days, but some individuals potentially reach peak viral shedding in under one day ^61^. The potential for an extremely rapid increase in viral load, which likely parallels shedding of infectious virus, means that in some cases, even daily testing might be insufficient to protect a community from someone who is newly infected.

Perhaps more importantly, the benefit of frequent testing of individuals without symptoms with RT-LAMP or other assays may be substantially undermined by risk disinhibition. When people are tested frequently, they may both underestimate their own risk of becoming infected in the interval between tests and overestimate the possibility that their similarly tested contacts are uninfected; anecdotal evidence of this phenomenon is perhaps most vividly seen in the September 26, 2020 White House Rose Garden reception for Justice Amy Coney Barrett, in which many attendees were photographed not wearing masks nor following guidelines for physical distancing ^62^. If infections among people without symptoms are rare (∼0.4% of tests in this study, when combining RT-LAMP and pooled qRT-PCR positives), but 10% of the tested population views testing as license for increased risk-tasking, is frequent testing of symptomless people a net positive? Appropriate messaging to the community is essential with any testing program to ensure the population understands the meaning of a test result. Such issues will require an optimization of messaging to mitigate the impact of risk disinhibition to the extent possible.

Ultimately, this study provides proof of concept and guidance for how decentralized rapid testing could be implemented in a mobile testing scenario, which may be especially useful in resource-limited settings. Despite the caveats presented above, we identified 12 SARS-CoV-2-positive individuals and likely prevented onward transmission from those individuals who otherwise would not know they were positive. Rapid tests, although less sensitive than qRT-PCR, have shorter turnaround times and could bridge the gap between SARS-CoV-2 surveillance and diagnostic testing. POC testing can be effective for identifying asymptomatic individuals but must be used in conjunction with appropriate messaging and other mitigation strategies to effectively reduce SARS-CoV-2 transmission.

## Data Availability

Data and protocols are available at https://openresearch.labkey.com/wiki/Coven/page.view?name=field-testing

## Acknowledgments

This work was made possible by financial support through NIH Rapid Acceleration of Diagnostics (3 U54 EB027690-02S1). Additional funding was provided in part by the office of the director, National Institutes of Health, under award number P51OD011106 to the Wisconsin National Primate Research Center (WNPRC), University of Wisconsin-Madison. This research was conducted in part at a facility constructed with support from Research Facilities Improvement Program grant numbers RR15459-01 and RR020141-01. The content is solely the responsibility of the authors and does not necessarily represent the official views of the National Institutes of Health. Additional funding was provided by the Wisconsin Alumni Research Foundation (WARF) COVID-19 Challenge. A.S.H. and C.M.C. have been supported by the National Institutes of Health National Research Service Award T32 AI007414. We are grateful for the assistance of both the University of Wisconsin Institutional Biosafety Committee and the Health Sciences Institutional Review Board at University of Wisconsin-Madison. We also acknowledge Christopher E. Mason, PhD Cornell-Weill Medical School and Alison Kriegel, PhD Medical College of Wisconsin as well as public employees from Racine, WI in their partnership to this project. We would like to thank Abigail Johnson, Abby Weaver, Abdel Daoud, Allison Eierman, Ben Boerigter, Clarissa Tjoanda, Julia Pulokas, Julie Chen, Ryan Anderson, WIll Vuyk, Jenny Lee, Max Bobholz, Sam Havlicek, Hunter Ries, Nicole Minerva, Emma Boehm, Elizabeth Brown, and Jayden Lee for their help preparing and organizing supplies for POC testing. David Beebe, Nate Grubaugh, and Kristian Andersen provided useful technical discussions. We thank Eli O’Connor for assistance with POC label generation and template preparation. We are grateful to the study volunteers and organizations that allowed us to evaluate this mobile testing strategy.

## Author Contributions

CMN, MDR, RWW, DMD, CGS, DHO, SLO contributed to assay development and optimization. DMD, MTM, RWW, CMN, MRS, AMW, MIB, KNF, MDR, LAH, OEH, RVM, CMC, SLO, MRR, TCF, TMP, EDS, LMS, EKN contributed to point of care testing and PCR confirmation. CMN, MDR, DMD, DHO contributed to data analysis, interpretation, and writing. JAK, DHO, SLO, HEB, TCF, MTM, AKH, LAH, CMC, KLH, CBB, KNF contributed to logistics and organization of point of care testing. CBB, KLH contributed to obtaining IRB and worked closely with the institutional biosafety committee on other regulatory responsibilities. MAA, ASH, WMR contributed to providing residual SARS-CoV-2 positive saliva samples and sample information from the University of Wisconsin Hospitals and Clinics. All authors contributed to editing the manuscript.

## Regulatory oversight

This work was performed under approved UW-Madison Health Sciences IRB #2020-0855 and #2020-1142.

