## Supplemental Table 1 for "Initial evaluation of a mobile SARS-CoV-2 RT-LAMP testing strategy"

Supplemental Table 1. POC RT-LAMP metrics

| Site | No. RT-LAMP assays | No. RT-LAMP-positive | No. potential false positive | No. qRT-PCR positive |
| --- | --- | --- | --- | --- |
| K-12_1 | 337 | 2 | 2 | 0 |
| K-12_2 | 170 | 0 | 0 | 0 |
| Research_1 | 524 | 4 | 1 | 3 |
| Research_2 | 2815 | 7 | 2 | 5 |
| Athletics | 858 | 8 | 3 | 4 |
| Total | 4704 | 21 | 8 | 12 |
